# Live virus neutralisation testing in convalescent patients and subjects vaccinated against 19A, 20B, 20I/501Y.V1 and 20H/501Y.V2 isolates of SARS-CoV-2

**DOI:** 10.1101/2021.05.11.21256578

**Authors:** Claudia Gonzalez, Carla Saade, Antonin Bal, Martine Valette, Kahina Saker, Bruno Lina, Laurence Josset, Mary-Anne Trabaud, Guillaume Thiery, Elisabeth Botelho-Nevers, Stéphane Paul, Paul Verhoeven, Thomas Bourlet, Sylvie Pillet, Florence Morfin, Sophie Trouillet-Assant, Bruno Pozzetto, on behalf of COVID-SER study group

## Abstract

**Background:** SARS-CoV-2 mutations appeared recently and can lead to conformational changes in the spike protein and probably induce modifications in antigenicity. In this study, we wanted to assess the neutralizing capacity of antibodies to prevent cell infection, using a live virus neutralisation test.

**Methods:** Sera samples were collected from different populations: two-dose vaccinated COVID-19-naïve healthcare workers (HCWs; Pfizer-BioNTech BNT161b2), 6-months post mild COVID-19 HCWs, and critical COVID-19 patients. We tested various clades such as 19A (initial one), 20B (B.1.1.241 lineage), 20I/501Y.V1 (B.1.1.7 lineage), and 20H/501Y.V2 (B.1.351 lineage).

**Results:** No significant difference was observed between the 20B and 19A isolates for HCWs with mild COVID-19 and critical patients. However, a significant decrease in neutralisation ability was found for 20I/501Y.V1 in comparison with 19A isolate for critical patients and HCWs 6-months post infection. Concerning 20H/501Y.V2, all populations had a significant reduction in neutralising antibody titres in comparison with the 19A isolate. Interestingly, a significant difference in neutralisation capacity was observed for vaccinated HCWs between the two variants whereas it was not significant for the convalescent groups.

**Conclusion:** Neutralisation capacity was slightly reduced for critical patients and HCWs 6-months post infection. No neutralisation escape could be feared concerning the two variants of concern in both populations. The reduced neutralising response observed towards the 20H/501Y.V2 in comparison with the 19A and 20I/501Y.V1 isolates in fully immunized subjects with the BNT162b2 vaccine is a striking finding of the study.

---

Several SARS-CoV-2 variants of concern (VOC) with mutations impacting notably the spike (S) protein have been detected recently [1]. These mutations can lead to conformational changes and probably induce modifications in antigenicity. Serological studies based on SARS-CoV-2 pseudotyped or chimeric viruses have been performed to measure the neutralisation activity of serum specimens of convalescent patients or subjects immunized by SARS-CoV-2 vaccines. Although these tests are easier to conduct, their ability to predict neutralising activity against authentic clinical viral isolates needs to be evaluated [2].

Herein we compared the ability of neutralising antibodies (NAb) directed against SARS-CoV-2 to prevent cell infection in different populations, using a live Virus Neutralisation Test (VNT) against SARS-CoV-2 isolates belonging to various clades: 19A, 20B (B.1.1.241 lineage), 20I/501Y.V1 (B.1.1.7 lineage) and 20H/501Y.V2 (B.1.351 lineage). VNT was performed as previously reported using an observed viral load of 100 to 500 50% Tissue Culture Infectious Doses (TCID_50_) for each isolate [3,4].

Serum specimens were collected from two-dose vaccinated COVID-19 naïve healthcare workers (HCWs; n=30) between two and four weeks after the administration of the Pfizer-BioNTech BNT162b2 vaccine (group 1), a subgroup of HCWs exhibiting significant NAb against 19A (50% plaque reduction neutralisation test (PRNT_50_) ranging from 20 to 240) 6-months after mild COVID-19 (n=29; group 2) [5], and critical COVID-19 patients sampled within one month after symptom onset (median [IQR]: 28 [22.5-33.5] days; n=25; group 3). All COVID-19 patients from groups 2 and 3 had been infected during the first wave of COVID-19 that occurred in France in March-April of 2020. Written informed consent was obtained from all HCWs; ethics approval was obtained from the national review board for biomedical research in April 2020 (Comité de Protection des Personnes Sud Méditerranée I, Marseille, France; ID RCB 2020-A00932-37), and the study was registered on ClinicalTrials.gov (NCT04341142). Concerning critical patients, this study was approved by the ethics committee of the university hospital of Saint-Etienne (reference number IRBN512020/CHUSTE).

No significant difference in median NAb titres was observed between the 19A isolate taken as reference and the 20B isolate that circulated during the second pandemic wave at the end of 2020 in Europe (Figure 1A). As previously reported between mild and severe patients [3], the three populations exhibited significantly different median levels of NAb for 19A isolate (Figure 1B); the same picture was observed for the two VOC (data not shown).

**Figure 1.**
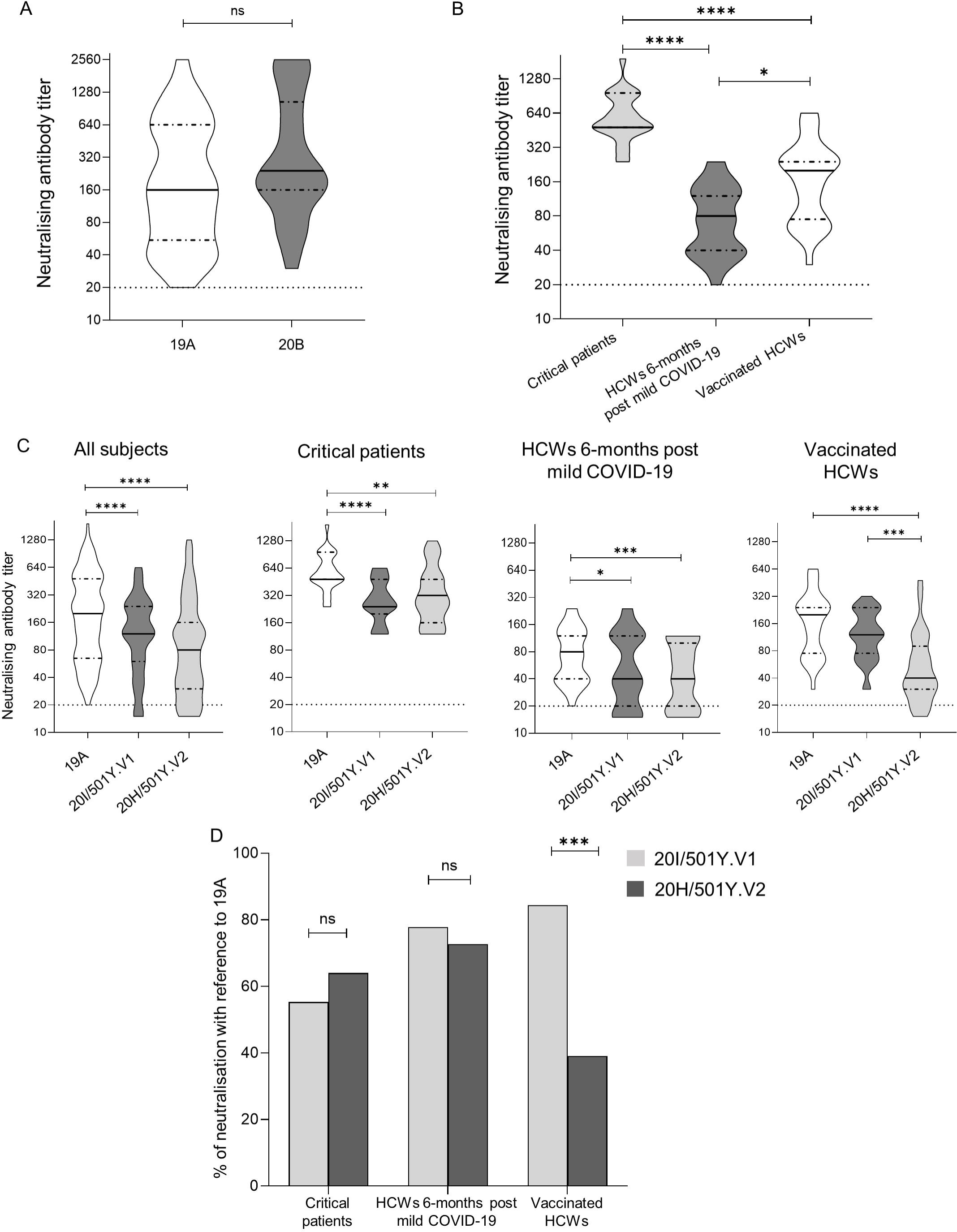
Neutralisation of living isolates of SARS-CoV-2 by convalescent sera from critical or mild patients with COVID-19 and by vaccine-elicited sera from subjects having received two doses of the BNT162b2 vaccine. *: p-value <0.05; **: p-value <0.01; ***: p-value <0.001; ****: p-value <0.0001; ns: non-significant. Each serum was tested in duplicate and the mean NAb was used for the analysis. Titres were transformed into log_2_ values for the calculation of mean NAb titres (a value of 0.5 was attributed by convention to negative samples). In panels A, B and C: dotted lines represent the detection threshold of 50% plaque reduction neutralisation test (PRNT_50_) ≥20 for neutralising antibody (NAb) titres; full lines represent median NAb titres and dash-dotted lines represent the 25% and 75% quartiles. (A) Violin plot presenting the neutralisation of 2 SARS-CoV-2 isolates belonging to the 19A and 20B clades by serum specimens obtained from subjects tested 6-months post mild COVID-19 (n=19) and critical patients (n=15); the Wilcoxon matched-pair signed rank test was used for comparisons. (B) Violin plot presenting the neutralisation of a SARS-CoV-2 isolate belonging to the 19A clade by serum specimens obtained from vaccinated subjects (n=30), 6-months post mild COVID-19 subjects (n=29) and critical patients (n=25); the Kruskal-Wallis test followed by Dunn’s multiple comparison test was used for comparisons. (C) Violin plots presenting the neutralisation of 3 SARS-CoV-2 isolates belonging to the 19A, 20I/501Y.V1 and 20H/501Y.V2 clades by the same serum specimens than in (B); the Friedman test followed by Dunn’s multiple comparison test was used for comparisons. (D) Capacity of neutralisation of the two tested VOC (20I/501Y.V1 and 20H/501Y.V2 clades) by the same serum specimens as that in (B) and (C) in comparison to a 19A isolate taken as reference; the Wilcoxon matched-pair signed rank test was used for comparisons.

By comparison to the 19A isolate, the median [IQR] fold reduction in NAb titres was 1.5 [1.2-1.7] and 3.5 [2.7-4.3] in group 1, 1.6 [1.3-1.8] and 1.9 [1.5-2.3] in group 2, and 2.3 [1.8-2.8] and 2.2 [1.5-2.8] in group 3, for the 20I/501Y.V1 and 20H/501Y.V2 VOC respectively (Figure 1C). Interestingly, the difference in median NAb titres was highly significant between the two variants (*p*<0.001) for vaccinated subjects (group 1) whereas it was not significant for the two other groups corresponding to naturally infected patients (Figure 1D).

As a whole, critical patients exhibited a strong neutralising response against all the tested strains; despite a slight reduction in NAb titres for both variants by comparison to the wild-type strain, no neutralisation escape could be feared against the two VOC due to the high titres of NAb.

The 6-months neutralising response of HCWs with mild COVID-19 was slightly reduced towards both variants by comparison to the wild type strain. By contrast to that reported herein, a study performed with pseudotype viruses on the same category of patients (6-months mild COVID-19 HCWs) found that the neutralising response was much lower against the 20H/501Y.V2 variant than against the 20I/501Y.V1 one [6], which would suggest an increased risk of neutralisation escape with the former strain.

Another striking finding of the present study is the reduced neutralising response observed towards the 20H/501Y.V2 variant in fully-immunised subjects with the BNT162b2 vaccine by comparison to the wild type and 20I/501Y.V1 variant. These results are in accordance with that observed for the same vaccine in numerous studies using pseudotype viruses [6-9] or authentic variant strains [10]. The neutralisation escape of the 20H/501Y.V2 variant in subjects having received either the BNT162b2 vaccine (mean fold reduction in NAb titre of 7.8) or the Oxford-AstraZeneca AZD1222 vaccine (mean fold reduction in NAb titres of 9) was shown to be mainly mediated by the synergistic effect of mutations K417N, E484K, and N501Y in the receptor binding domain [11].

Although the present study was performed using a live VNT, it focused only on the humoral response and other experiments are needed to assess the overall immune process including T-cell immune response. Another limitation of the study regards the huge differences between the three tested populations in terms of time post-infection or immunisation, which restrains the interpretation of the differences in results between the three groups.

In conclusion, the relative good conservation of the neutralising activity of sera from the three populations tested in this study against the two variants 20I/501Y.V1 and 20H/501Y.V2 is encouraging towards a putative reinfection by these strains. Long-term monitoring of the NAb response together with that of the specific cellular response will be needed to confirm these favourable findings.

## Data Availability

GISAID accession numbers:

EPI_ISL_1707038

EPI_ISL_1707039

EPI_ISL_1707040

EPI_ISL_768828

## Data availability statement

GISAID accession numbers:

EPI_ISL_1707038

EPI_ISL_1707039

EPI_ISL_1707040

EPI_ISL_768828

## Acknowledgements

We are indebted to all the personnel of the occupational health and medicine department of Hospices Civils de Lyon who contributed to the collection of samples, especially Virginie Pitiot, Fanny Joubert and PMO team. Human biological samples and associated data were obtained from NeuroBioTec (CRB HCL, Lyon France, Biobank BB-0033-00046). We thank Karima Brahami and all members of the clinical research and innovation department for their reactivity (DRS, Hospices Civils de Lyon). All members of Saint-Etienne Hospital who contributed to the collection of samples are acknowledged.

